# Employer Requirements and COVID-19 Vaccination and Attitudes among Healthcare Personnel in the U.S.: Findings from National Immunization Survey Adult COVID Module, August – September 2021

**DOI:** 10.1101/2022.03.14.22271847

**Authors:** James T. Lee, S. Sean Hu, Tianyi Zhou, Kim Bonner, Jennifer L. Kriss, Elisabeth Wilhelm, Rosalind J. Carter, Carissa Holmes, Marie A. de Perio, Peng-jun Lu, Kimberly H. Nguyen, Noel T. Brewer, James A. Singleton

## Abstract

**Introduction:** Employer vaccination requirements have been used to increase vaccination uptake among healthcare personnel (HCP). In summer 2021, HCP were the group most likely to have employer requirements for COVID-19 vaccinations as healthcare facilities led the implementation of such requirements. This study examined the association between employer requirements and HCP’s COVID-19 vaccination status and attitudes about the vaccine.

**Methods:** Participants were a national representative sample of United States (US) adults who completed the National Immunization Survey Adult COVID Module (NIS-ACM) during August–September 2021. Respondents were asked about COVID-19 vaccination and intent, requirements for vaccination, place of work, attitudes surrounding vaccinations, and sociodemographic variables. This analysis focused on HCP respondents. We first calculated the weighted proportion reporting COVID-19 vaccination for HCP by sociodemographic variables. Then we computed unadjusted and adjusted prevalence ratios for vaccination coverage and key indicators on vaccine attitudes, comparing HCP based on individual self-report of vaccination requirements.

**Results:** Of 12,875 HCP respondents, 41.5% reported COVID-19 vaccination employer requirements. Among HCP with vaccination requirements, 90.5% had been vaccinated against COVID-19, as compared to 73.3% of HCP without vaccination requirements—a pattern consistent across sociodemographic groups. Notably, the greatest differences in uptake between HCP with and without employee requirements were seen in sociodemographic subgroups with the lowest vaccination uptake, e.g., HCP aged 18–29 years, HCP with high school or less education, HCP living below poverty, and uninsured HCP. In every sociodemographic subgroup examined, vaccine uptake was more equitable among HCP with vaccination requirements than in HCP without. Finally, HCP with vaccination requirements were also more likely to express confidence in the vaccine’s safety (68.3% vs. 60.1%) and importance (89.6% vs 79.6%).

**Conclusion:** In a large national US sample, employer requirements were associated with higher and more equitable HCP vaccination uptake across all sociodemographic groups examined. Our findings suggest that employer requirements can contribute to improving COVID-19 vaccination coverage, similar to patterns seen for other vaccines.

## 1. Introduction

To protect patients and staff, US healthcare facilities have adopted vaccination requirements for a wide range of vaccine-preventable diseases, including measles (Parker-Fiebelkorn, 2014), hepatitis B (Lindley, 2007), pertussis (Lu, 2014), varicella (Lindley, 2011), and seasonal influenza (Greene, 2018). Employer requirements for seasonal influenza vaccines are associated with higher coverage (Black, 2018), and have been found to accelerate institution-wide coverage (Miller, 2011; Norwalk, 2013), as well as sustaining high coverage over the course of a decade (Blank, 2020).

US healthcare personnel (HCP) were among the first groups to receive access to COVID-19 vaccinations in December 2020, prioritized ahead of other essential workers and most seniors (Dooling, 2020, Goodman NYT). However, by July 2021, 1-dose vaccination uptake among surveyed HCP had plateaued at around 75% (NCIRD COVID Vax Views). For example, in North Carolina, 14 state-operated health facilities with about 10,000 employees undertook extensive staff education, individualized counseling, and on-site vaccinations over a six-month period. Yet, COVID-19 vaccination coverage was 75% at the end of June 2021 (NC DHHS, 2021). Further studies suggest that vaccination uptake was inequitable among HCP, with lower vaccination coverage among lower paid occupations and in facilities located in areas of high social vulnerability (Lee, 2021; Gharpure, 2021). In this context, individual health facilities began to put in place vaccination requirements for staff (Miller, 2021; Ritter, 2021). By November, 2021 more than 12 US states and the District of Columbia (Kaiser Family Foundation, 2021) enforced COVID-19 vaccination requirements for HCP. On November 4, 2021, the US Centers for Medicare and Medicaid Services (CMS) issued an emergency interim final rule requiring all Medicare- and Medicaid-certified providers to establish COVID-19 vaccination requirements for staff (CMS, 2021).

While COVID-19 vaccination employer requirements have become more common, the assessment of such policies remains limited (WH Report, 2021). Policymakers, employers, and consumers continue to express concern that employer requirements may be ineffective, harden vaccine hesitancy, or even lead to paradoxical decreases in vaccine uptake. In this study, we describe self-reported employer vaccination requirements and its association with COVID-19 vaccination uptake, and key attitudes surrounding vaccination beliefs among US HCP.

## 2. Methods

### 2.1 Participants and procedures

The National Immunization Survey Adult COVID Module (NIS-ACM) is a nationally representative survey with approximately 60,000 adult respondents (aged 18 years and older) monthly. The NIS-ACM conducts telephone interviews from a random-digit-dialed sample of cell telephone numbers stratified by state and the District of Columbia as well as Puerto Rico and the U.S. Virgin Islands. Our study focused analysis on 12,875 HCP respondents interviewed during August 1–September 25, 2021 from 50 states and the District of Columbia excluding Puerto Rico and the U.S. Virgin Islands. This is a subset of 91,771 total respondents during this period with an overall survey response rate of 20.5% during August 1–28 and 20.9% during August 29–September 25. The Centers for Disease Control and Prevention (CDC) determined that the NIS-ACM constitutes public health surveillance. NIS-ACM was conducted consistent with applicable federal law and CDC policy.^1^

### 2.2 Measures

Respondents were classified as HCP based on their answers to the questions “Are you a frontline or essential worker according to your state region?” and “In what location or setting do you currently work?” Respondents were classified as HCP if they answered 1) “yes” or 2) “don’t know” to the first question and selected “healthcare (e.g., hospital, doctor, dentist or mental health specialist office, outpatient facility, long-term care, home healthcare, pharmacy, medical laboratory)” for their location and setting of work. These questions were designed to correspond to categories of essential workers recommended for prioritization (Dooling, 2021). Respondents were asked “Does your work or school require you to get a COVID-19 vaccine?” to assess employer requirements for vaccinations.

Respondents were categorized as vaccinated if they reported having received one or more doses of COVID-19 vaccines. For unvaccinated respondents, vaccination intent was assessed (“How likely are you to get a COVID-19 vaccine? Would you say you would definitely get a vaccine, probably get a vaccine, probably not get a vaccine, definitely not get a vaccine, or are not sure?”); we further categorized those answering “not sure” or “probably get a vaccine” into a “more reachable” group and those answering “probably not get a vaccine” or “definitely not get a vaccine” into a “reluctant” group.

The NIS-ACM included questions assessing the behavioral and social drivers of vaccination (Brewer, 2021), described in detail and available online (NCIRD NIS-ACM, 2021). For this study, we analyzed questions assessing how respondents think and feel about vaccinations – specifically questions around the perceived importance of the vaccine (“How important do you think getting a COVID-19 vaccine is to protect yourself against COVID-19?”), safety (“How safe do you think a COVID-19 vaccine is for you?”), and anticipated regret for not vaccinating (“If I do not get (had not gotten) a COVID-19 vaccine, I will regret (would have regretted) it”).

Respondents were asked about their sex, race/ethnicity, age, education, household income, health insurance status, and zip code or city of residence. Urbanicity, as defined by metropolitan statistical area (MSA) classification (MSA principal city, MSA non-principal city, and non-MSA), was determined based on household reported city and county of residence (OMB, 2021). Household income was categorized relative to US Census Bureau’s 2020 poverty threshold and at the level of $75,000 (US Census Bureau).

### 2.3 Statistical Analysis

The data were weighted to represent the non-institutionalized U.S. population and mitigate possible bias that can result from an incomplete sample frame (exclusion of households with no phone service or only landline telephones) or non-response. Survey weights were also calibrated to jurisdiction-level vaccine administration data (stratified by age group and sex) reported to CDC as of mid-month for each approximate monthly analytic data file (CDC COVID Data Tracker). T-tests were used to identify differences in prevalence of vaccination uptake from multivariable logistic between each response level and the reference group for each sociodemographic variable (D’Agostino, 1988). T-tests were then used to test for differences in the unadjusted prevalence ratio (PR) from multivariable logistic to determine the differences in prevalence of vaccination uptake comparing HCP with and without an employer requirement for each social demographic variable and each vaccine attitude variable. Furthermore, T-tests were also used to test for differences in the adjusted prevalence ratio (aPR) from multivariable logistic to determine the differences in prevalence of vaccination uptake comparing HCP with and without an employer requirement for each vaccine attitude variable; prevalence estimates were adjusted for age, sex, race/ethnicity, and education level. Estimates, along with 95% confidence intervals (CIs), were calculated using SAS-callable SUDAAN (Research Triangle Institute, Research Triangle Park, NC, version 11.0.1) to account for the complex survey design. All differences were tested using two-tailed t-tests with a significance level set at α = 0.05.

## 3. Results

### 3.1 Vaccination Status and Employer Requirements

Among all HCP respondents, 80.3% (95% Confidence Interval: 78.8%–81.7%) reported receiving ≥1 dose of COVID-19 vaccine (Table 1). In total, 41.5% (95% CI: 40.0%–43.1%) of HCP reported employer requirements for COVID-19 vaccination – a percentage that increased from 35.0% in August to 48.3% in September. Among HCP reporting vaccination requirements, 90.5% reported receiving ≥1 dose of COVID-19 vaccine, as compared to 73.3% among HCP not reporting vaccination requirements – a crude prevalence ratio of 1.24 (95% CI: 1.19–1.28; Figure 1, Table 1). We also found substantial geographic variations in the prevalence of employer COVID-19 vaccination requirements and vaccination uptake (Figure 2).

**Table 1.**
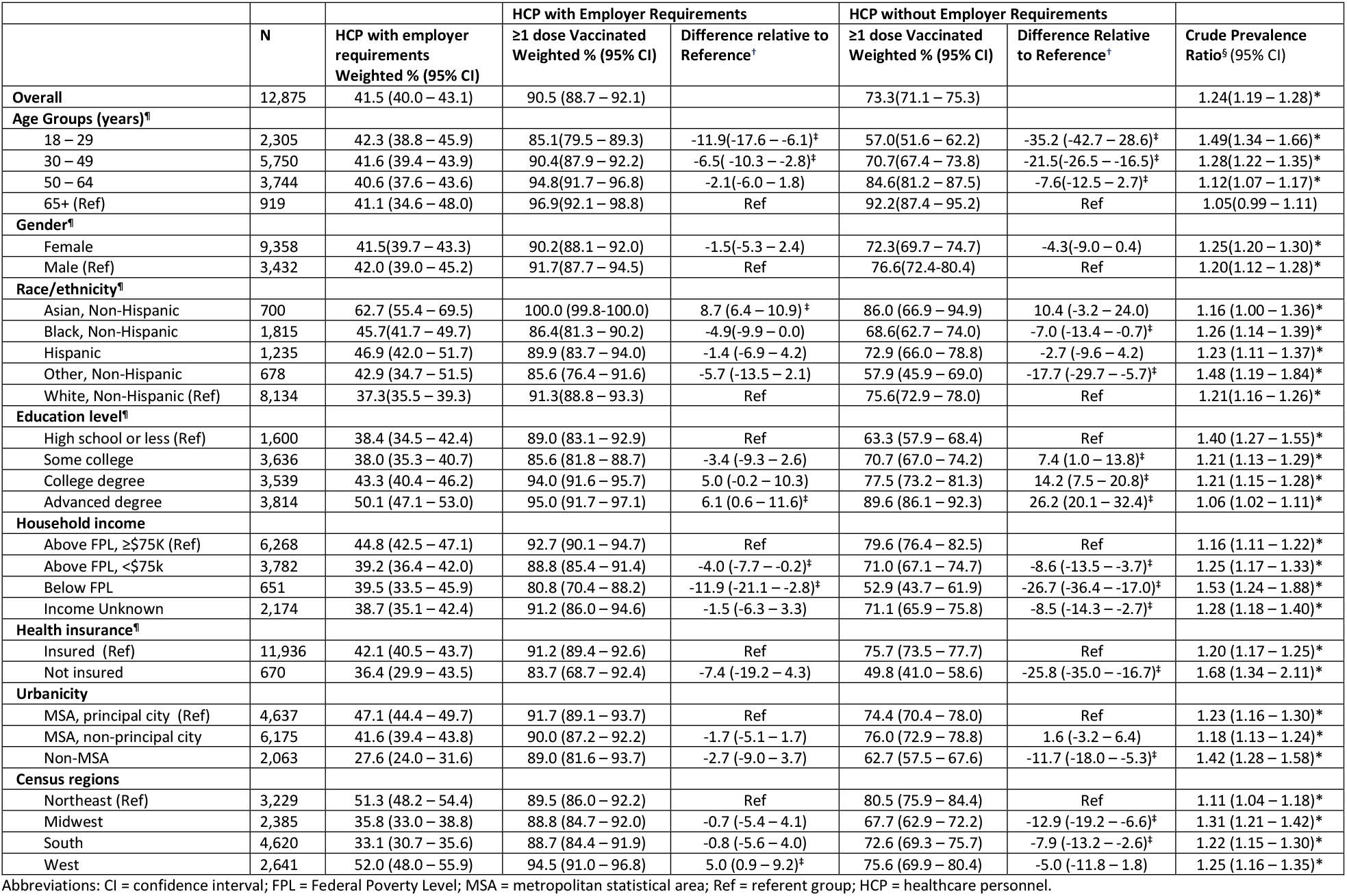
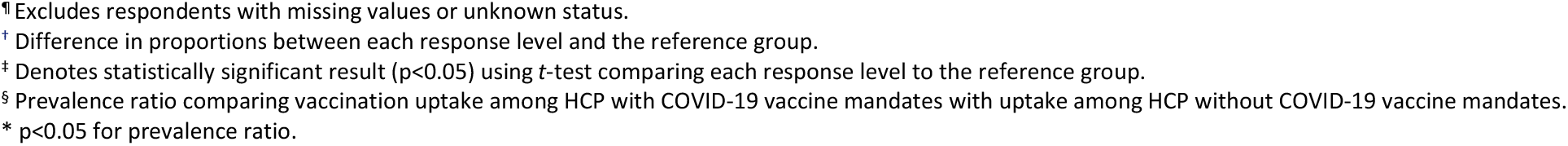
COVID–19 vaccination status, by employer vaccination requirement and sociodemographic characteristics, NIS-ACM, August 1 – September 25 2021.

|  |  |  | HCP with Employer Requirements |  | HCP without Employer Requirements |  |  |
| --- | --- | --- | --- | --- | --- | --- | --- |
|  | N | HCP with employer requirements<br>Weighted % (95% CI) | ≥1 dose Vaccinated<br>Weighted % (95% CI) | Difference relative to<br>Reference<sup>†</sup> | ≥1 dose Vaccinated<br>Weighted % (95% CI) | Difference Relative<br>to Reference<sup>†</sup> | Crude Prevalence<br>Ratio<sup>§</sup> (95% CI) |
| **Overall** | 12,875 | 41.5 (40.0 – 43.1) | 90.5 (88.7 – 92.1) |  | 73.3(71.1 – 75.3) |  | 1.24(1.19 – 1.28)* |
| **Age Groups (years)<sup>¶</sup>** |  |  |  |  |  |  |  |
| 18 – 29 | 2,305 | 42.3 (38.8 – 45.9) | 85.1(79.5 – 89.3) | -11.9(-17.6 – -6.1)<sup>‡</sup> | 57.0(51.6 – 62.2) | -35.2 (-42.7 – 28.6)<sup>‡</sup> | 1.49(1.34 – 1.66)* |
| 30 – 49 | 5,750 | 41.6 (39.4 – 43.9) | 90.4(87.9 – 92.2) | -6.5(-10.3 – -2.8)<sup>‡</sup> | 70.7(67.4 – 73.8) | -21.5(-26.5 – -16.5)<sup>‡</sup> | 1.28(1.22 – 1.35)* |
| 50 – 64 | 3,744 | 40.6 (37.6 – 43.6) | 94.8(91.7 – 96.8) | -2.1(-6.0 – 1.8) | 84.6(81.2 – 87.5) | -7.6(-12.5 – 2.7)<sup>‡</sup> | 1.12(1.07 – 1.17)* |
| 65+ (Ref) | 919 | 41.1 (34.6 – 48.0) | 96.9(92.1 – 98.8) | Ref | 92.2(87.4 – 95.2) | Ref | 1.05(0.99 – 1.11) |
| **Gender<sup>¶</sup>** |  |  |  |  |  |  |  |
| Female | 9,358 | 41.5(39.7 – 43.3) | 90.2(88.1 – 92.0) | -1.5(-5.3 – 2.4) | 72.3(69.7 – 74.7) | -4.3(-9.0 – 0.4) | 1.25(1.20 – 1.30)* |
| Male (Ref) | 3,432 | 42.0 (39.0 – 45.2) | 91.7(87.7 – 94.5) | Ref | 76.6(72.4-80.4) | Ref | 1.20(1.12 – 1.28)* |
| **Race/ethnicity<sup>¶</sup>** |  |  |  |  |  |  |  |
| Asian, Non-Hispanic | 700 | 62.7 (55.4 – 69.5) | 100.0 (99.8-100.0) | 8.7 (6.4 – 10.9)<sup>‡</sup> | 86.0 (66.9 – 94.9) | 10.4 (-3.2 – 24.0) | 1.16 (1.00 – 1.36)* |
| Black, Non-Hispanic | 1,815 | 45.7(41.7 – 49.7) | 86.4(81.3 – 90.2) | -4.9(-9.9 – 0.0) | 68.6(62.7 – 74.0) | -7.0 (-13.4 – -0.7)<sup>‡</sup> | 1.26 (1.14 – 1.39)* |
| Hispanic | 1,235 | 46.9 (42.0 – 51.7) | 89.9 (83.7 – 94.0) | -1.4 (-6.9 – 4.2) | 72.9 (66.0 – 78.8) | -2.7 (-9.6 – 4.2) | 1.23 (1.11 – 1.37)* |
| Other, Non-Hispanic | 678 | 42.9 (34.7 – 51.5) | 85.6 (76.4 – 91.6) | -5.7 (-13.5 – 2.1) | 57.9 (45.9 – 69.0) | -17.7 (-29.7 – -5.7)<sup>‡</sup> | 1.48 (1.19 – 1.84)* |
| White, Non-Hispanic (Ref) | 8,134 | 37.3(35.5 – 39.3) | 91.3(88.8 – 93.3) | Ref | 75.6(72.9 – 78.0) | Ref | 1.21(1.16 – 1.26)* |
| **Education level<sup>¶</sup>** |  |  |  |  |  |  |  |
| High school or less (Ref) | 1,600 | 38.4 (34.5 – 42.4) | 89.0 (83.1 – 92.9) | Ref | 63.3 (57.9 – 68.4) | Ref | 1.40 (1.27 – 1.55)* |
| Some college | 3,636 | 38.0 (35.3 – 40.7) | 85.6 (81.8 – 88.7) | -3.4 (-9.3 – 2.6) | 70.7 (67.0 – 74.2) | 7.4 (1.0 – 13.8)<sup>‡</sup> | 1.21 (1.13 – 1.29)* |
| College degree | 3,539 | 43.3 (40.4 – 46.2) | 94.0 (91.6 – 95.7) | 5.0 (-0.2 – 10.3) | 77.5 (73.2 – 81.3) | 14.2 (7.5 – 20.8)<sup>‡</sup> | 1.21 (1.15 – 1.28)* |
| Advanced degree | 3,814 | 50.1 (47.1 – 53.0) | 95.0 (91.7 – 97.1) | 6.1 (0.6 – 11.6)<sup>‡</sup> | 89.6 (86.1 – 92.3) | 26.2 (20.1 – 32.4)<sup>‡</sup> | 1.06 (1.02 – 1.11)* |
| **Household income** |  |  |  |  |  |  |  |
| Above FPL, ≥\$75K (Ref) | 6,268 | 44.8 (42.5 – 47.1) | 92.7 (90.1 – 94.7) | Ref | 79.6 (76.4 – 82.5) | Ref | 1.16 (1.11 – 1.22)* |
| Above FPL, <\$75k | 3,782 | 39.2 (36.4 – 42.0) | 88.8 (85.4 – 91.4) | -4.0 (-7.7 – -0.2)<sup>‡</sup> | 71.0 (67.1 – 74.7) | -8.6 (-13.5 – -3.7)<sup>‡</sup> | 1.25 (1.17 – 1.33)* |
| Below FPL | 651 | 39.5 (33.5 – 45.9) | 80.8 (70.4 – 88.2) | -11.9 (-21.1 – -2.8)<sup>‡</sup> | 52.9 (43.7 – 61.9) | -26.7 (-36.4 – -17.0)<sup>‡</sup> | 1.53 (1.24 – 1.88)* |
| Income Unknown | 2,174 | 38.7 (35.1 – 42.4) | 91.2 (86.0 – 94.6) | -1.5 (-6.3 – 3.3) | 71.1 (65.9 – 75.8) | -8.5 (-14.3 – -2.7)<sup>‡</sup> | 1.28 (1.18 – 1.40)* |
| **Health insurance<sup>¶</sup>** |  |  |  |  |  |  |  |
| Insured (Ref) | 11,936 | 42.1 (40.5 – 43.7) | 91.2 (89.4 – 92.6) | Ref | 75.7 (73.5 – 77.7) | Ref | 1.20 (1.17 – 1.25)* |
| Not insured | 670 | 36.4 (29.9 – 43.5) | 83.7 (68.7 – 92.4) | -7.4 (-19.2 – 4.3) | 49.8 (41.0 – 58.6) | -25.8 (-35.0 – -16.7)<sup>‡</sup> | 1.68 (1.34 – 2.11)* |
| **Urbanicity** |  |  |  |  |  |  |  |
| MSA, principal city (Ref) | 4,637 | 47.1 (44.4 – 49.7) | 91.7 (89.1 – 93.7) | Ref | 74.4 (70.4 – 78.0) | Ref | 1.23 (1.16 – 1.30)* |
| MSA, non-principal city | 6,175 | 41.6 (39.4 – 43.8) | 90.0 (87.2 – 92.2) | -1.7 (-5.1 – 1.7) | 76.0 (72.9 – 78.8) | 1.6 (-3.2 – 6.4) | 1.18 (1.13 – 1.24)* |
| Non-MSA | 2,063 | 27.6 (24.0 – 31.6) | 89.0 (81.6 – 93.7) | -2.7 (-9.0 – 3.7) | 62.7 (57.5 – 67.6) | -11.7 (-18.0 – -5.3)<sup>‡</sup> | 1.42 (1.28 – 1.58)* |
| **Census regions** |  |  |  |  |  |  |  |
| Northeast (Ref) | 3,229 | 51.3 (48.2 – 54.4) | 89.5 (86.0 – 92.2) | Ref | 80.5 (75.9 – 84.4) | Ref | 1.11 (1.04 – 1.18)* |
| Midwest | 2,385 | 35.8 (33.0 – 38.8) | 88.8 (84.7 – 92.0) | -0.7 (-5.4 – 4.1) | 67.7 (62.9 – 72.2) | -12.9 (-19.2 – -6.6)<sup>‡</sup> | 1.31 (1.21 – 1.42)* |
| South | 4,620 | 33.1 (30.7 – 35.6) | 88.7 (84.4 – 91.9) | -0.8 (-5.6 – 4.0) | 72.6 (69.3 – 75.7) | -7.9 (-13.2 – -2.6)<sup>‡</sup> | 1.22 (1.15 – 1.30)* |
| West | 2,641 | 52.0 (48.0 – 55.9) | 94.5 (91.0 – 96.8) | 5.0 (0.9 – 9.2)<sup>‡</sup> | 75.6 (69.9 – 80.4) | -5.0 (-11.8 – 1.8) | 1.25 (1.16 – 1.35)* |

Abbreviations: CI = confidence interval; FPL = Federal Poverty Level; MSA = metropolitan statistical area; Ref = referent group; HCP = healthcare personnel.
<sup>¶</sup> Excludes respondents with missing values or unknown status.
<sup>†</sup> Difference in proportions between each response level and the reference group.
<sup>‡</sup> Denotes statistically significant result ( $p < 0.05$ ) using *t*-test comparing each response level to the reference group.
<sup>§</sup> Prevalence ratio comparing vaccination uptake among HCP with COVID-19 vaccine mandates with uptake among HCP without COVID-19 vaccine mandates.
\* $p < 0.05$ for prevalence ratio.

**Table 2.** COVID-19 Vaccination Status and Attitudes among Healthcare Personnel, by COVID-19 Vaccine Requirement, National Immunization Survey-Adult COVID Module, United States, August 1 – September 25 2021.

| Variable | HCP with Employer Requirements % (95% CI) | HCP without Employer Requirements % (95%CI) | Crude Prevalence Ratio (95%CI) | Adjusted Prevalence Ratio<sup>§</sup> (95%CI) |
| --- | --- | --- | --- | --- |
| **≥1 dose Vaccinated** | 90.5 (88.7-92.1) | 73.3 (71.1-75.3) | 1.24 (1.19-1.28)* | 1.21 (1.17-1.25)* |
| **Unvaccinated, “Definitely get a vaccine”** | 1.9 (1.4-2.7) | 1.8 (1.3-2.4) | 1.09 (0.68-1.75) | 1.19 (0.73-1.96) |
| **Unvaccinated, more reachable** | 3.3 (2.3-4.5) | 8.7 (7.3-10.4) | 0.37 (0.26-0.54)* | 0.39 (0.26-0.58)* |
| **Unvaccinated, reluctant** | 4.3 (3.2-5.7) | 16.2 (14.5-18.1) | 0.27 (0.20-0.36)* | 0.27 (0.20-0.38)* |
| **Vaccine is “very” or “somewhat” important to protect yourself** | 89.6 (87.9-91.1) | 79.6 (77.6-81.5) | 1.13 (1.09-1.16)* | 1.10 (1.07-1.13)* |
| **Vaccine is “completely” or “very safe”** | 68.3 (65.8-70.7) | 60.1 (57.8-62.3) | 1.14 (1.08-1.20)* | 1.11 (1.05-1.16)* |
| **“Very” or “Strongly agree” with anticipated regret statement** | 62.9 (60.5-65.2) | 50.9 (48.7-53.1) | 1.23 (1.17-1.31)* | 1.18 (1.11-1.25)* |

Abbreviations: CI = confidence interval.
<sup>§</sup> Prevalence ratio comparing vaccination uptake and attitudes among health care personnel with COVID-19 vaccine requirement with rates among those without COVID-19 vaccine requirement, adjusted for age, sex, race/ethnicity, and education level..
\* p<0.05 for prevalence ratio.

**Figure 1.**
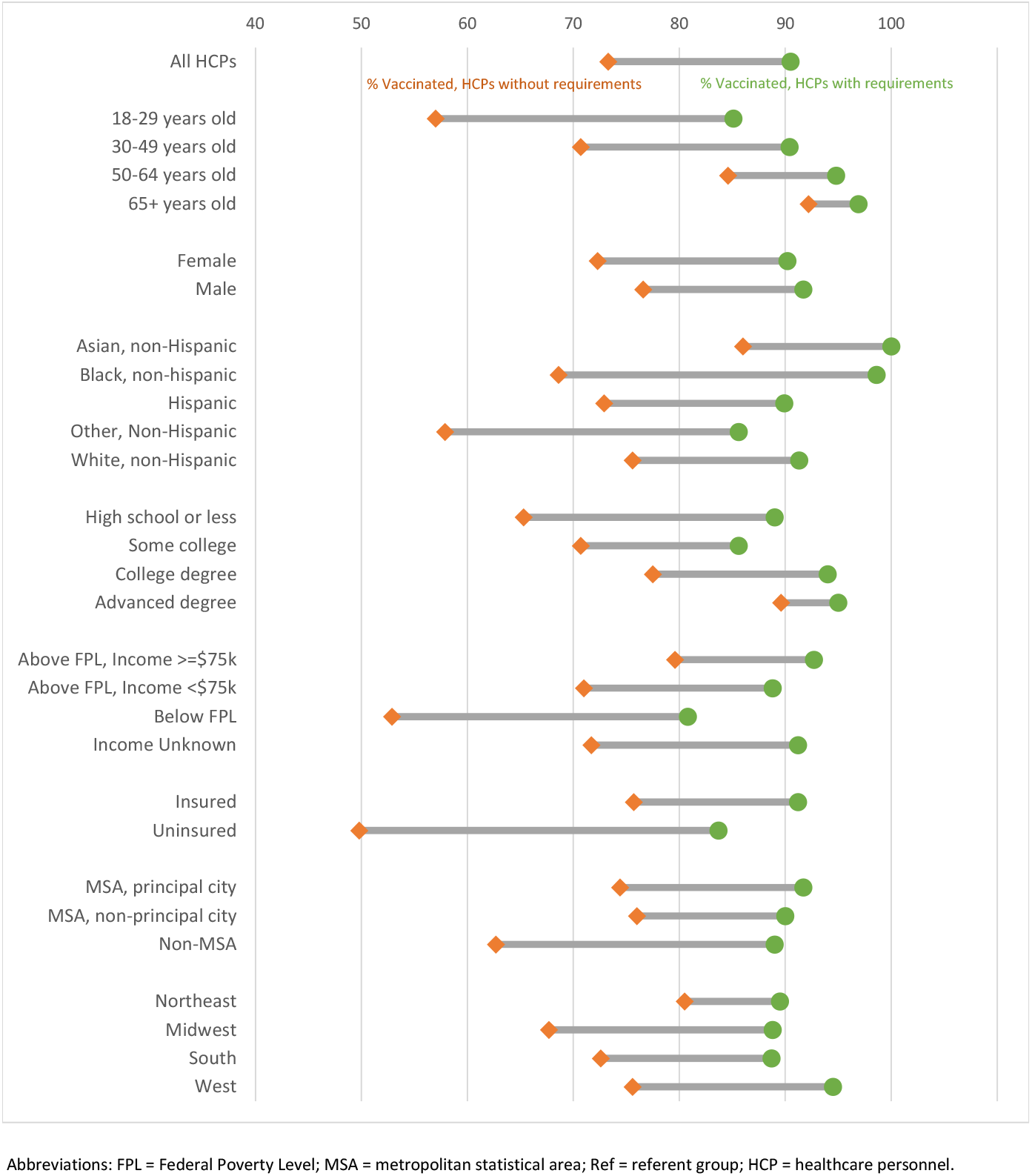
COVID–19 vaccination coverage of healthcare personnel, by employer vaccination requirement status and sociodemographic characteristics, NIS–ACM, August 1 – September 25, 2021. Abbreviations: FPL = Federal Poverty Level; MSA = metropolitan statistical area; Ref = referent group; HCP = healthcare personnel.

**Figure 2.**
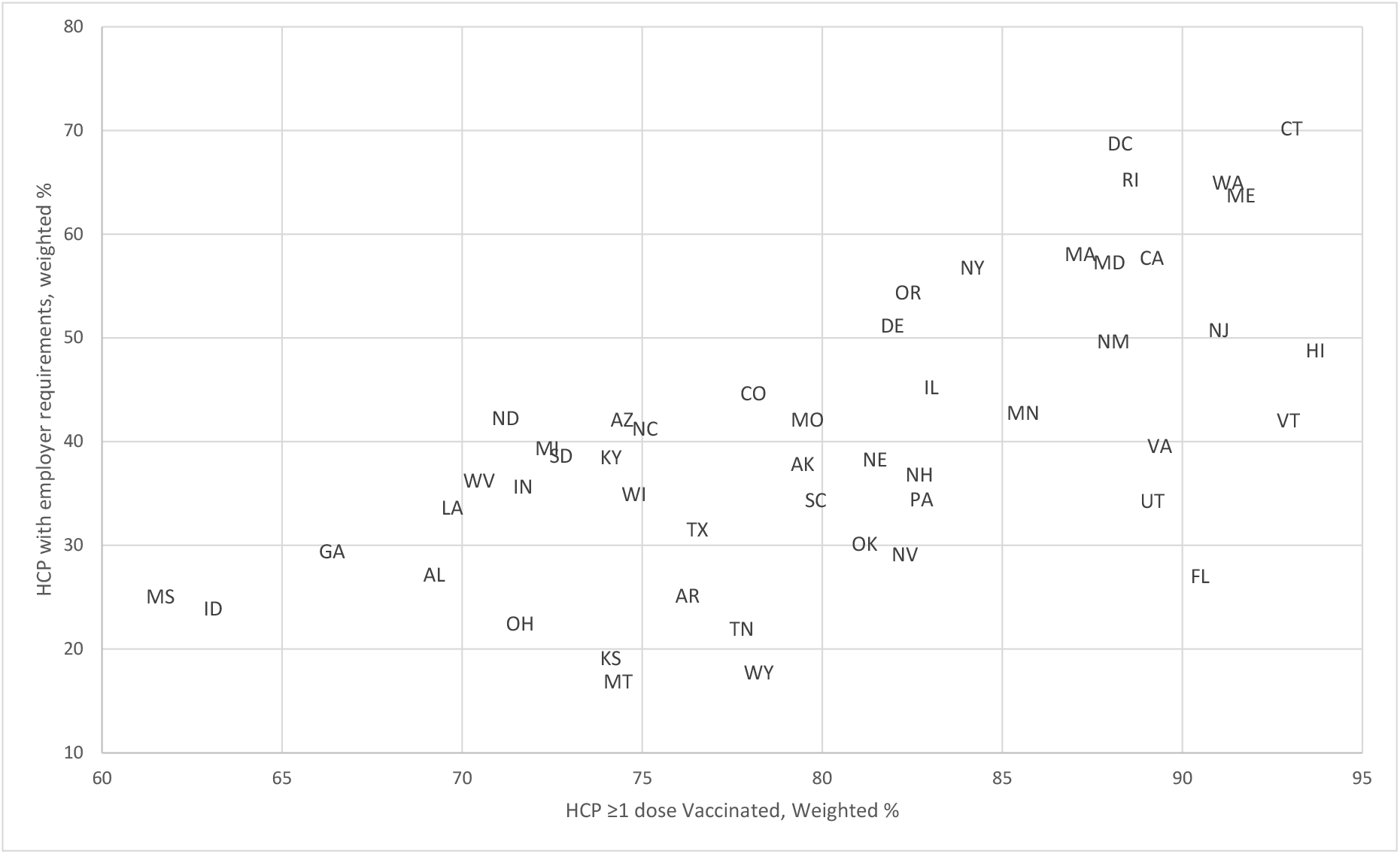
COVID-19 Vaccination Uptake and Employer Requirement among Healthcare Personnel by State, National Immunization Survey-Adult COVID Module, United States, August 1 – September 25 2021.

HCP who faced employer requirements were more likely to report COVID-19 vaccinations in every sociodemographic subgroup. Within each sociodemographic subgroup, HCP demonstrated similar disparities to US adults overall in COVID-19 vaccination uptake. Lower COVID-19 vaccination coverage was seen among younger HCP, HCP with lower education attainment or lower incomes, uninsured HCP, and HCP residing in non-MSAs or in the US Midwest (Figure 1; Table 1).

For every demographic group, vaccine uptake was more equitable among HCP reporting employer vaccination requirements. Within every sociodemographic subgroup, the difference in vaccination uptake associated with employer requirements was greatest in the groups with the lowest COVID-19 vaccination coverage (Figure 1). Examples include crude prevalence ratios of 1.49 (95% CI: 1.34–1.66) for HCP 18-29 years old, 1.53 (95% CI: 1.24–1.88) among HCP living below the Federal Poverty Line, 1.68 (95% CI: 1.34–2.11) for uninsured HCP, 1.42 (95% CI: 1.28–1.58) for HCP living in Non-MSAs, and 1.31 (95% CI: 1.21 – 1.42) for HCP living in the US Midwest (Table 1).

### 3.2 Key Attitudes about COVID-19 Vaccination

For unvaccinated HCP, employer requirements were associated with differences in vaccination intent. HCP with vaccination requirements had fewer unvaccinated HCP in two categories – 4.3% versus 16.2% in the reluctant group (aPR = 0.27, 95% CI: 0.20–0.38); 3.3% versus 8.7% in the “more reachable” group (aPR of 0.39; 95% CI: 0.26– 0.58). A similar percentage of HCP with and without requirements reported that they “definitely plan to get vaccinated.” The reluctant group had the greatest difference between those reporting employer requirements and those not reporting requirements. Approximately 43% of unvaccinated HCP with reporting requirements were categorized as reluctant compared with nearly 61% of unvaccinated HCP without requirements.

HCP reporting vaccination requirements were more likely to say that the vaccine was somewhat or very important to protect themselves than HCP without vaccination requirements (89.6% versus 79.6%; Apr = 1.10, 95% CI: 1.07 – 1.13). Relative to HCP without requirements, HCP with vaccination requirements were also more likely to say that the COVID-19 vaccine was very or completely safe (68.3% vs 60.1%; aPR 1.11, 95% CI = 1.05–1.16) and either strongly agree or very strongly agree with the statement that “If I do not get a COVID-19 vaccine, I will regret it” (62.9% vs 50.9%; aPR = 1.18, 95% CI: 1.11–1.25).

## 4. Discussion

Employer COVID-19 vaccination requirements have become more common for HCP in the United States and expanding through CMS requirements. Compared to HCP without employer requirements, HCP with employer requirements had 17.2 percentage points higher ≥1 dose COVID-19 vaccination coverage, a difference that persisted even after adjusting for sociodemographic characteristics of HCP surveyed. Notably, we found the largest requirement versus no-requirement difference in sociodemographic groups with the lowest coverage – some of these groups have experienced disproportionate COVID-19 morbidity and mortality (e.g., African Americans and Latino, lower income groups) while other groups have persistently lower vaccine uptake despite the widespread availability of vaccines (e.g., those aged 18-29 years). Even for employees who reported employer requirements, 9.4% remained unvaccinated – this may be the result of the time between the announcements and enforcement of employer requirements, the presence of religious and medical exemptions, degree of enforcement in employer policies, as well as limitations in survey self-report.

Vaccination requirements were associated with greater confidence in the importance and safety of COVID-19 vaccines. NIS-ACM’s cross-sectional design precludes conclusions on the directionality of vaccination attitudes and vaccination status – it is possible that vaccination requirements were imposed on HCP with greater confidence in COVID-19 vaccines. However, it is also possible that receipt of COVID-19 vaccines induces greater confidence in its safety and importance, a pattern of post-hoc attitudinal changes seen in other vaccinations and health behavior (Levy-Bruhl, 2019; Hall, 2018).

Employer vaccination requirements may also have a role in converting vaccination intent into action. For HCP who report employer requirements, the percentage of vaccinated HCP approximates the percentage who said that the vaccination is important to protect themselves – at 90.5% and 89.6% respectively. For HCP without employer requirements, while 79.6% said that the COVID-19 vaccine is important to protect themselves, only 73.3% are vaccinated.

Finally, employer requirements for HCP vaccinations have come months after the initial availability of COVID-19 vaccines in the US, after extensive post-authorization monitoring and reporting, and when most eligible Americans were vaccinated against COVID-19. During this period, over 65% of Americans reported that many or almost all of their friends and family have been vaccinated (CDC COVID VaxViews) – a social norm that is likely more prevalent in healthcare institutions. HCP are also more likely to have access to workplace vaccinations, consultations with other HCP as trusted sources of information, as well as encountering strong provider recommendations – critical strategies in building vaccine confidence prior to implementing vaccination requirements (CDC COVID-19 Vaccination Field Guide).

The findings in this report are subject to limitations. First, NIS-ACM does not include adults living in institutionalized settings and phoneless or landline-only households, which might introduce the possibility for selection bias. However, their exclusion would not be expected to introduce any major bias, because only 2.3% of US adults reported having no telephone service or using landline only during July-December 2020 (Blumberg, 2021). Second, the low response rate can increase the potential for bias if respondents and nonrespondents differ systematically, even after adjusting for nonresponse. Estimates of COVID-19 vaccination coverage might differ from vaccine administration and other data reported elsewhere (CDC COVID-19 Vaccinations in the US, 2021). Third, these data are based on self-report; therefore, they are subject to reporting and recall bias. Calibration of survey weights to the vaccine administration data likely mitigated at least some of the possible bias from nonresponse and misclassification of self-reported COVID-19 vaccination status. Fourth, NIS-ACM does not collect occupational classes of HCP, though the variable on education level can approximate such dynamics. Finally, the question on requirements included both work and school requirements. It is possible that some HCP reported vaccinations requirements were school requirements, although this number is likely to be very limited.

## 5. Conclusion

Our findings demonstrate that employer requirements for HCP COVID-19 vaccinations are associated with 1) higher vaccination uptake; 2) smaller vaccination disparities within every examined sociodemographic subgroup, driven by increase in vaccination uptake in the most vulnerable and least vaccinated groups; and 3) greater confidence in the importance and safety of the vaccine among respondent HCP. While the cross-sectional nature of our survey precludes causal inferences, these findings suggest employer vaccination requirements are an instrument to improve vaccination uptake, equity, and confidence.

## Data Availability

Data produced in the present work are contained in the manuscript and also available at https://www.cdc.gov/vaccines/imz-managers/coverage/covidvaxview/interactive/adults.html

https://data.cdc.gov/Vaccinations/National-Immunization-Survey-Adult-COVID-Module-NI/akkj-j5ru/data

## Footnotes

1 § See e.g., 45 C.F.R. part 46.102(l)(2), 21 C.F.R. part 56; 42 U.S.C. §241(d); 5 U.S.C. §552a; 44 U.S.C. §3501 et seq.

## Notes

### Competing Interest Statement

None of the authors have no conflicts of interest relevant to this article to disclose. Noel Brewer has served as a paid consultant for CDC, WHO, Merck, and Novartis; none of the other of the authors have financial relationships relevant to this article to disclose.

### Funding Statement

This study did not receive any funding.

### Author Declarations

The Institutional Review Board of the United States Centers for Disease Control and Prevention (Atlanta, Georgia, United States) waived ethical review approval for this work as non-research public health surveillance in accordance to 45 C.F.R. part 46.102 (l) (2). This activity was reviewed under protocol number 0900f3eb81ccfa24.

## REFERENCES

1. Black CL. Influenza Vaccination Coverage Among Health Care Personnel — United States, 2017– 18 Influenza Season. MMWR Morb Mortal Wkly Rep 2018;67. https://doi.org/10.15585/mmwr.mm6738a2

2. Blank C, Gemeinhart N, Dunagan WC, Babcock HM. Mandatory employee vaccination as a strategy for early and comprehensive health care personnel immunization coverage: Experience from 10 influenza seasons. American Journal of Infection Control 2020;48:1133–8. https://doi.org/10.1016/j.ajic.2020.01.015.

3. Blumberg S, Luke JV. Wireless Substitution: Early Release of Estimates From the National Health Interview Survey, January-June 2020. National Center for Health Statistics; 2021. https://doi.org/10.15620/cdc:100855.

4. Brewer NT. What Works to Increase Vaccination Uptake. Acad Pediatr 2021;21:S9–16. https://doi.org/10.1016/j.acap.2021.01.017.

5. Centers for Disease Control and Prevention. COVID-19 Vaccination Field Guide: 12 COVID-19 Vaccination Strategies for Your Community. Centers for Disease Control and Prevention 2021. https://www.cdc.gov/vaccines/covid-19/vaccinate-with-confidence/community.html (accessed November 17, 2021).

6. Centers for Disease Control and Prevention. COVID-19 Vaccinations in the United States. Centers for Disease Control and Prevention 2021. https://covid.cdc.gov/covid-data-tracker/#vaccinations_vacc-total-admin-rate-total (accessed November 17, 2021).

7. Centers for Medicare & Medicaid Services (CMS). Medicare and Medicaid Programs; Omnibus COVID-19 Health Care Staff Vaccination. Federal Register 2021. https://www.federalregister.gov/documents/2021/11/05/2021-23831/medicare-and-medicaid-programs-omnibus-covid-19-health-care-staff-vaccination (accessed November 8, 2021).

8. Community Preventative Services Task Force (CPSTF). Vaccination Programs: Requirements for Child Care, School, and College Attendance. The Guide to Community Preventive Services (The Community Guide) 2016. https://www.thecommunityguide.org/findings/vaccination-programs-requirements-child-care-school-and-college-attendance (accessed November 15, 2021).

9. D’agostino RB, Chase W, Belanger A. The Appropriateness of Some Common Procedures for Testing the Equality of Two Independent Binomial Populations. The American Statistician 1988;42:198–202. https://doi.org/10.1080/00031305.1988.10475563.

10. Dooling K. The Advisory Committee on Immunization Practices’ Interim Recommendation for Allocating Initial Supplies of COVID-19 Vaccine — United States, 2020. MMWR Morb Mortal Wkly Rep 2020;69. https://doi.org/10.15585/mmwr.mm6949e1.

11. Dooling K. The Advisory Committee on Immunization Practices’ Updated Interim Recommendation for Allocation of COVID-19 Vaccine — United States, December 2020. MMWR Morb Mortal Wkly Rep 2021;69. https://doi.org/10.15585/mmwr.mm695152e2.

12. Gharpure R, Yi SH, Li R, Jacobs Slifka KM, Tippins A, Jaffe A, et al. COVID-19 Vaccine Uptake Among Residents and Staff Members of Assisted Living and Residential Care Communities— Pharmacy Partnership for Long-Term Care Program, December 2020–April 2021. Journal of the American Medical Directors Association 2021;22:2016-2020.e2. https://doi.org/10.1016/j.jamda.2021.08.015.

13. Goodman JD, Ferré-Sadurní L. ‘Big Fight’ Breaks Out Over Which Interest Groups Get Vaccine First. The New York Times 2020.

14. Greene MT, Fowler KE, Ratz D, Krein SL, Bradley SF, Saint S. Changes in Influenza Vaccination Requirements for Health Care Personnel in US Hospitals. JAMA Network Open 2018;1:e180143. https://doi.org/10.1001/jamanetworkopen.2018.0143.

15. Hall MG, Marteau TM, Sunstein CR, Ribisl KM, Noar SM, Orlan EN, et al. Public support for pictorial warnings on cigarette packs: An experimental study of US smokers. J Behav Med 2018;41:398–405. https://doi.org/10.1007/s10865-018-9910-2.

16. Kaiser Family Foundation. COVID-19 Vaccine Mandates, as of November 10, 2021. KFF 2021. https://www.kff.org/report-section/state-covid-19-data-and-policy-actions-policy-actions/ (accessed November 11, 2021).

17. Lee JT, Althomsons SP, Wu H, Budnitz DS, Kalayil EJ, Lindley MC, et al. Disparities in COVID-19 Vaccination Coverage Among Health Care Personnel Working in Long-Term Care Facilities, by Job Category, National Healthcare Safety Network — United States, March 2021. MMWR Morb Mortal Wkly Rep 2021;70:1036–9. https://doi.org/10.15585/mmwr.mm7030a2.

18. Lévy-Bruhl D, Fonteneau L, Vaux S, Barret A-S, Antona D, Bonmarin I, et al. Assessment of the impact of the extension of vaccination mandates on vaccine coverage after 1 year, France, 2019. Euro Surveill 2019;24:1900301. https://doi.org/10.2807/1560-7917.ES.2019.24.26.1900301.

19. Lindley MC, Horlick GA, Shefer AM, Shaw FE, Gorji M. Assessing State Immunization Requirements for Healthcare Workers and Patients. American Journal of Preventive Medicine 2007;32:459–65. https://doi.org/10.1016/j.amepre.2007.02.009.

20. Lindley MC, Lorick SA, Spinner JR, Krull AR, Mootrey GT, Ahmed F, et al. Student Vaccination Requirements of U.S. Health Professional Schools: A Survey. Ann Intern Med 2011;154:391–400. https://doi.org/10.7326/0003-4819-154-6-201103150-00004.

21. Lu P, Graitcer SB, O’Halloran A, Liang JL. Tetanus, diphtheria and acellular pertussis (Tdap) vaccination among healthcare personnel—United States, 2011. Vaccine 2014;32:572–8. https://doi.org/10.1016/j.vaccine.2013.11.077.

22. Miller BL, Ahmed F, Lindley MC, Wortley PM. Increases in vaccination coverage of healthcare personnel following institutional requirements for influenza vaccination: A national survey of US hospitals. Vaccine 2011;29:9398–403. https://doi.org/10.1016/j.vaccine.2011.09.047.

23. Miller SM, Phillips RA, Schwartz RL, Sostman HD, Hackett C, Boom ML. How to Develop a Covid-19 Employee Vaccination Policy. Harvard Business Review 2021.

24. National Center for Immunization and Respiratory Diseases. COVIDVaxView Interactive! | CDC 2021. https://www.cdc.gov/vaccines/imz-managers/coverage/covidvaxview/interactive.html (accessed November 11, 2021).

25. National Center for Immunization and Respiratory Diseases. NIS Adult COVID Module (NIS-ACM) Hard Copy Questionnaire: Q3/2021 n.d. https://www.cdc.gov/vaccines/imz-managers/nis/downloads/NIS-ACM-Questionnaire-Q3-2021.pdf.

26. North Carolina Department of Health and Human Services. Nearly All Staff in State Operated Healthcare Facilities Fully Vaccinated 2021. https://www.ncdhhs.gov/news/press-releases/2021/10/11/nearly-all-staff-state-operated-healthcare-facilities-fully-vaccinated (accessed November 8, 2021).

27. Nowalk MP, Lin CJ, Raymund M, Bialor J, Zimmerman RK. Impact of hospital policies on health care workers’ influenza vaccination rates. American Journal of Infection Control 2013;41:697– 701. https://doi.org/10.1016/j.ajic.2012.11.011.

28. Office of Management and Budget (OMB). 2010 Standards for Delineating Metropolitan and Micropolitan Statistical Areas. Federal Register 2010. https://www.federalregister.gov/documents/2010/06/28/2010-15605/2010-standards-for-delineating-metropolitan-and-micropolitan-statistical-areas (accessed November 17, 2021).

29. Parker Fiebelkorn A, Seward JF, Orenstein WA. A global perspective of vaccination of healthcare personnel against measles: Systematic review. Vaccine 2014;32:4823–39. https://doi.org/10.1016/j.vaccine.2013.11.005.

30. Ritter AZ, Kelly J, Kent RM, Howard P, Theil R, Cavanaugh P, et al. Implementation of a Coronavirus Disease 2019 Vaccination Condition of Employment in a Community Nursing Home. Journal of the American Medical Directors Association 2021;22:1998–2002. https://doi.org/10.1016/j.jamda.2021.07.035.

31. Shapiro GK, Kaufman J, Brewer NT, Wiley K, Menning L, Leask J, et al. A critical review of measures of childhood vaccine confidence. Current Opinion in Immunology 2021;71:34–45. https://doi.org/10.1016/j.coi.2021.04.002.

32. The White House. WHITE HOUSE REPORT: Vaccination Requirements Are Helping Vaccinate More People, Protect Americans from COVID-19, and Strengthen the Economy 2021. https://www.whitehouse.gov/wp-content/uploads/2021/10/Vaccination-Requirements-Report.pdf.

33. US Census Bureau. How the Census Bureau Measures Poverty. CensusGov n.d. https://www.census.gov/topics/income-poverty/poverty/guidance/poverty-measures.html (accessed November 17, 2021).

